# Factors that influenced the course of the mpox outbreak in the Netherlands, 2022-2023

**DOI:** 10.1101/2023.10.09.23296426

**Authors:** Manon R. Haverkate, Inge J.M. Willemstein, Catharina E. van Ewijk, Philippe C.G. Adam, Susan J. Lanooij, Petra Jonker-Jorna, Colette van Bokhoven, Gini G.C. van Rijckevorsel, Elske Hoornenborg, Silke David, Liesbeth Mollema, Margreet J. te Wierik, Jente Lange, Eelco Franz, Hester E. de Melker, Eline L.M. Op de Coul, Susan J.M. Hahné

**Affiliations:** Centre for Infectious Disease Control, National Institute for Public Health and the Environment, PO Box 1, 3720 BA Bilthoven, the Netherlands; European Programme for Intervention Epidemiology Training (EPIET), European Centre for Disease Prevention and Control (ECDC), 171 83 Stockholm, Sweden; Institute for Prevention and Social Research, Utrecht, the Netherlands; Centre for Social Research in Health, University of New South Wales Sydney, NSW 2052, Sydney, Australia; Public Health Service Hollands Noorden, PO box 9276, 1800 GG Alkmaar, the Netherlands; Public Health Service Gelderland-Zuid, PO box 1120, 6501 BC Nijmegen, the Netherlands; Public Health Service Amsterdam, PO box 2200, 1000 CE Amsterdam, the Netherlands

## Abstract

**Background:** In 2022-2023 a global outbreak of mpox affected mostly gay, bisexual and other men having sex with men (GBMSM). Outbreak control in the Netherlands included isolation, quarantine, post-exposure prophylaxis vaccination, and primary preventive vaccination (PPV). We describe the course of the outbreak, the vaccination programme, vaccine effectiveness (VE) of full vaccination against symptomatic disease, and trends in behaviour to generate hypotheses about factors that influenced the outbreak’s decline.

**Methods:** Observational study. Data on notified cases, number of PPV invitations, and PPV doses administered were collected from public health services. PPV uptake and coverage were calculated. Monthly trends in behavioural data of GBMSM visiting Sexual Health Centres (SHC) were analysed for all consultations in 2022. VE was estimated using the screening method.

**Findings:** A total of 1,266 mpox cases were reported until August 1, 2023. The outbreak peaked early July 2022 and sharply declined afterwards. PPV started July 25, 2023; in total 29,851 doses were administered. 45.8% received at least one dose, 35.4% was fully vaccinated. Estimated VE was 68.2% (95% CI 4.3%-89.5%). In our analyses of behavioural data, we did not observe an evident decrease in high-risk behaviour.

**Interpretation:** It is unlikely that PPV was a driver of the outbreak’s decline, as incidence started to decline well before the start of the PPV programme. The possible impact of behavioural change could not be demonstrated with the available indicators. We hypothesise that infection-induced immunity in high-risk groups contributed most to the decline.

**Funding:** Dutch Ministry of Health, Welfare and Sport.

## Background

Since the first reported human case of mpox (caused by monkeypox virus (MPXV)) in 1970 in the Democratic Republic of the Congo, endemic transmission of mpox was for decades only reported from African countries. Sporadic travel-associated cases were described outside of the African continent,^1,2^ and in 2003 an outbreak in the US was caused by imported mammals.^3–5^ In 2022 a global outbreak of mpox occurred, affecting mostly gay, bisexual and other men who have sex with men (GBMSM).^1,6,7^ Up to August 1, 2023 there were over 88,000 confirmed cases worldwide in 113 countries.^8^ The first patient in the Netherlands was diagnosed May 20, 2022 and the outbreak peaked in the beginning of July. The first 1,000 cases in the outbreak of mpox in the Netherlands until August 8, 2022 have been described by van Ewijk et al.^9^ Since then, the incidence continued to decrease. Up to August 1, 2023 an additional 266 persons with laboratory confirmed mpox have been reported to the Dutch National Institute for Public Health and the Environment (RIVM), resulting in a total of 1,266 confirmed mpox cases. In 2023, only 6 infections were diagnosed, of which the most recently diagnosed person had a date of symptom onset of May 26, 2023.

Outbreak control in the Netherlands included isolation of persons with mpox, and quarantine (later replaced by lifestyle recommendations) and post-exposure prophylaxis (PEP) vaccination with Modified Vaccinia Ankara-Bavarian Nordic (MVA-BN; also known as Imvanex or Jynneos), a third-generation smallpox vaccine,^10^ for high- and medium-risk contacts of all individuals diagnosed with mpox. Also, a primary preventive vaccination (PPV) programme with MVA-BN started on July 25, 2022, targeting those at highest risk of infection.^11^ Some variability in starting dates existed between different regions, but all regions started before August 11, 2022. Vaccination against smallpox was ceased in 1976 in the Netherlands.^12^ It is estimated that at the start of the 2022 mpox outbreak, 57%^9^ to 68%^13^ of the Dutch population did not have protective antibodies against Orthopox viruses. GBMSM and transgender persons were alerted on behaviours that would put them at higher risk of mpox and preventive measures to mitigate the risk through a targeted national communication campaign.^14^ Here, we describe the mpox outbreak in the Netherlands, the design and implementation of the vaccination programme, and trends in behavioural data to generate hypotheses about the factors that may have contributed to the decline of the outbreak in the Netherlands. Furthermore, we aim to estimate the vaccine effectiveness of MVA-BN against symptomatic mpox disease.

## Methods

### Data collection

All individuals diagnosed with mpox were reported to the RIVM in an online reporting system as part of the mandatory notification (see Supplementary Information).^15^ For all individuals diagnosed with mpox, information was collected on, amongst others, demographics, sexual orientation, potential source(s) of infection and results of contact tracing. A detailed description of the variables collected can be found in van Ewijk et al.^9^

The following groups were eligible for PPV: 1a. GBMSM and transgender persons using HIV-pre-exposure prophylaxis (PrEP) via Sexual Health Centres (SHC); 1b. GBMSM and transgender persons on the waiting list for the HIV-PrEP-pilot via SHC; 1c. GBMSM and transgender persons using HIV-PrEP via their general practitioner (GP); 2. GBMSM and transgender persons living with HIV at high-risk for mpox infection; 3. Other GBMSM and transgender persons at high-risk for mpox infection.^11^ All Public Health Services (PHS) identified persons eligible for PPV, and personally invited them. All vaccinations were given subcutaneously. The estimated numbers of PPV invitations sent out per indication group and all administered PPV doses were registered by all PHS in a dedicated online reporting system. For vaccinated persons that did not give consent to share their vaccination information with the RIVM, only the following anonymised data were collected: year of birth in three categories (<1985, 1985-1994, >1994), childhood smallpox vaccination yes/no, vaccination number (1^st^ or 2^nd^), week of vaccination, and PHS location. For persons that gave consent to share pseudonymised data, this was supplemented with: unique identifier, year of birth, gender, indication group, immunocompromised (yes/no), received PEP vaccination (yes/no), date of PEP vaccination, and date of PPV. Administered PEP vaccinations were not registered on a national level. Details on PEP vaccination, the PPV programme and selection of persons eligible for PPV can be found in the Supplementary Information.

In the Netherlands, data regarding all visits to SHC are available in a national surveillance database (SOAP) of the RIVM. This database contains pseudonymised data from all consultations, including behavioural data. Consultations can be divided into regular SHC consultations and HIV-PrEP-pilot consultations. Persons participating in the HIV-PrEP-pilot are regularly tested for STIs at SHC (three-monthly), independently of their sexual behaviour. Monitoring behaviour in this group will therefore give a more accurate representation of possible changes in behaviour, compared to persons who consult a SHC because of symptoms or possible risk incurred. Therefore, only HIV-PrEP consultations of GBMSM and transgender persons (including non-binary persons) were included. As persons could have multiple consultations in 2022, analyses were done on consultation level. Variables extracted from the SOAP database used as proxies for behaviour with high risk for mpox infection were number of sex partners, group sex (yes/no; defined as multiple sex partners at once or over a short period), chemsex (yes/no; defined as using drugs [cocaine, XTC/MDMA/Speed, Heroin, Crystal Meth, Mephedrone, 3-MMC, 4-MEC, 4-FA, GHB/GBL and ketamine] before or during sex), condomless anal sex (always condomless vs. sometimes/never; receptive and/or insertive) and sex work (yes/no). All behavioural variables are self-reported behaviour in the past six months. Furthermore, number of tests and percentage positivity for chlamydia, gonorrhoea and syphilis were also analysed.

Data on notifications were included until August 1, 2023, data on vaccinated persons were included until April 30, 2023; and behavioural data were collected from all SHC consultations in 2022.

### Data analyses

For individuals diagnosed with mpox, we assumed people born ≥1976 in the Netherlands or ≥1980 in another country did not receive a childhood smallpox vaccination.

Mpox vaccine uptake was defined as the proportion of invited people receiving at least one dose. Mpox vaccination coverage was defined as the proportion of invited people being fully vaccinated. A person was considered fully vaccinated if they had received two MVA-BN vaccinations; or if they received one MVA-BN vaccination and a childhood smallpox vaccination and were not reported to be immunocompromised. For the mpox vaccinees we had no information on country of birth. We assumed that people born <1976 with an unknown childhood smallpox vaccination status did receive this vaccination as a child, while childhood smallpox vaccination status was set to ‘No’ for everybody born ≥1980. For those born between 1976 and 1980, smallpox vaccination status was used as reported. If it was unknown if a person was immunocompromised, we assumed they were not. For both the analyses of the uptake and vaccination coverage, vaccinations administered to people who were not in the predefined indication groups were excluded. Uptake and vaccination coverage were calculated for all invitees, and by indication group.

Vaccine effectiveness (VE) against laboratory confirmed symptomatic mpox disease was estimated for those who were fully vaccinated using the screening method as described by Farrington.^16^ This method compares the vaccination coverage in the cases with the coverage in the population they arose from, with the VE calculated as 1 – odds of vaccination in cases/odds of vaccination in the population. Cases were included in the analyses if they were male at birth, indicated they had sex with men, and if their vaccination status was known. For these analyses, persons were considered fully vaccinated using the definition described before, plus their last vaccination should be at least 14 days before symptom onset to allow time for vaccine-induced protection.^17^ Also, at least one of the vaccinations received should be a PPV, to make the cases comparable to the population where the vaccination coverage was calculated from. Cases that had received their last vaccination less than 14 days before onset of symptoms were regarded incompletely vaccinated and were included in the total number of cases, but not in the completely vaccinated cases. The proportion of fully vaccinated among the cases was calculated per week. The vaccination coverage in the population (i.e., the proportion of fully vaccinated persons of those invited for PPV) was calculated per week as well. Here, also a 14-day delay was considered to allow time for vaccine-induced protection. Increasing vaccination coverage was accounted for by fitting a logistic regression model with the logit of the proportion of the population vaccinated as offset.^16^ To reduce bias introduced by the small sample size, Firth correction was applied to the logistic regression model. As the number of mpox infections dropped dramatically by the end of 2022, with 0 persons to include in December, analyses were done for August up to and including November.

To describe trends in behaviour among GBMSM and transgender persons, the numbers and proportions of consultations in which each sexual behaviour variable was reported, and the number of tests and positivity rates for chlamydia, gonorrhoea and infectious syphilis were plotted per month. To assess a possible significant decrease in risk behaviour (p<0.05), we compared the proportions in June, July and August (outbreak months) to May (pre-outbreak month) using Chi square tests. For non-normally distributed continuous variables the Mann-Whitney U test was used. All analyses were done in R version 4.2.0 and Excel version 2022.

## Results

### Epidemiology

Of all individuals diagnosed with mpox in the Netherlands, nearly all (99%; 1,247/1,265, for 1 person sex was unknown) were male at birth, of whom 94% (1,171/1,247) reported to have sex with men. Of those for whom information about gender identity was known, 98% identified as a man (1,076/1,098). The median age was 37 years (IQR 31-45). Of those eligible for childhood smallpox vaccination (born <1976 in the Netherlands or <1980 in another country) and with complete smallpox vaccination information, only 62% (168/273) reported to have received this vaccination in the past. Of those with information about PEP vaccination status for mpox, 4% (52/1,171) reported to have received PEP, but only in two instances it was given at least 14 days before the date of symptom onset. Of those with a known PPV status, 7% (56/768) reported to have received PPV at any time during the outbreak. In 12 instances the last vaccination was given at least 14 days before the date of symptom onset. 51% (640/1,266) of the individuals diagnosed with mpox originated from the Amsterdam region. Demographical, epidemiological and clinical characteristics of the 266 more recently diagnosed persons are comparable to the first 1,000 cases. For further details on the characteristics of these cases, we refer to van Ewijk et al.^9^

### Vaccination

In total, 39,657 invitations for vaccination were sent out by the 25 PHS in the Netherlands. Overall, 29,851 PPV doses were administered between July 25, 2022 and April 5, 2023. Consent to share vaccination information with the RIVM was given for 26,993 vaccinations (90.4%). In total, 17,792 first vaccinations were given, and 11,846 second. 213 vaccinations had an unknown ranking number. These first and second PPV doses were administered to 18,684 unique persons, predominantly identifying as man. Background characteristics of the vaccinated persons can be found in Table 1.

**Table 1.**
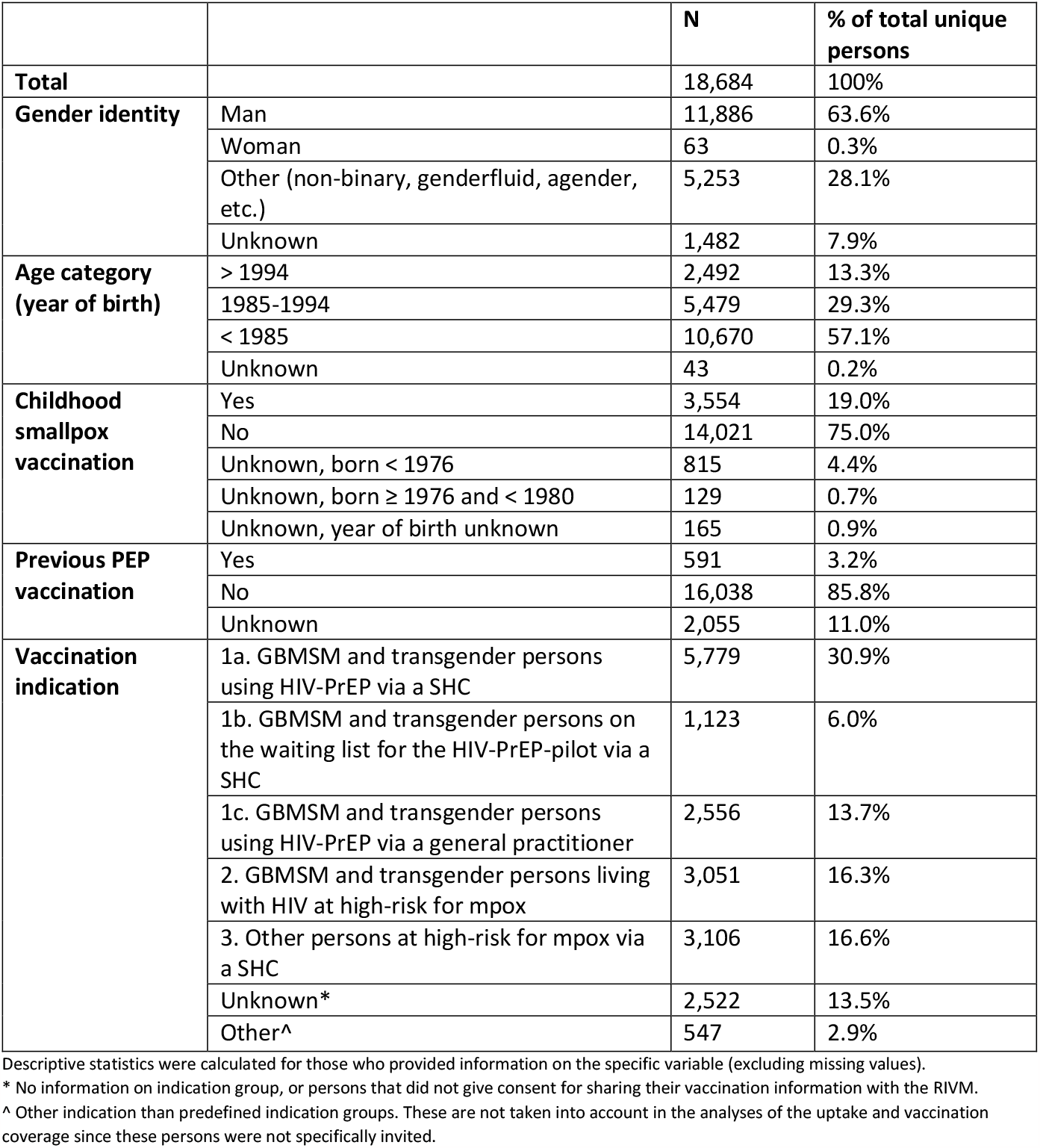
Background characteristics of mpox vaccinated persons (PPV) in the Netherlands; July 25, 2022-April 5, 2023.

The overall uptake of vaccination (at least one dose received) was 45.8% (18,157/39,657) with 35.4% (14,048/39,657) being fully vaccinated by April 30, 2023, with considerable variation between the different indication groups (Table 2). The highest uptake and vaccination coverage were found in GBMSM and transgender persons using HIV-PrEP, either via a SHC (indication group 1a) or GP (indication group 1c). The vaccination coverage in relation to the epidemic curve can be found in Figure 1.

**Table 2.** Invited persons, uptake and vaccination coverage per indication group.

| **Vaccination indication** | **Invited persons\*** | **Uptake^** | **Vaccination coverage#** |
| --- | --- | --- | --- |
| 1a. GBMSM and transgender persons using HIV-PrEP via a SHC | 8,702 | 66.4%<br>(5,779/8,702) | 55.4%<br>(4,825/8,702) |
| 1b. GBMSM and transgender persons on the waiting list for the HIV-PrEP-pilot via a SHC | 2,776 | 40.5%<br>(1,123/2,776) | 33.5% (929/2,776) |
| 1c. GBMSM and transgender persons using HIV-PrEP via a general practitioner | 4,811 | 53.1%<br>(2,556/4,811) | 42.4%<br>(2,041/4,811) |
| 2. GBMSM and transgender persons living with HIV at high-risk for mpox | 9,858 | 30.9%<br>(3,051/9,858) | 16.7%<br>(1,648/9,858) |
| 3. Other persons at high-risk for mpox via a SHC | 13,484 | 23.0%<br>(3,106/13,484) | 18.2%<br>(2,448/13,484) |

\* For 26 invitations it was unknown which indication group the person belonged to
^ Uptake: proportion of invited persons receiving at least one dose MVA-BN

**Figure 1.**
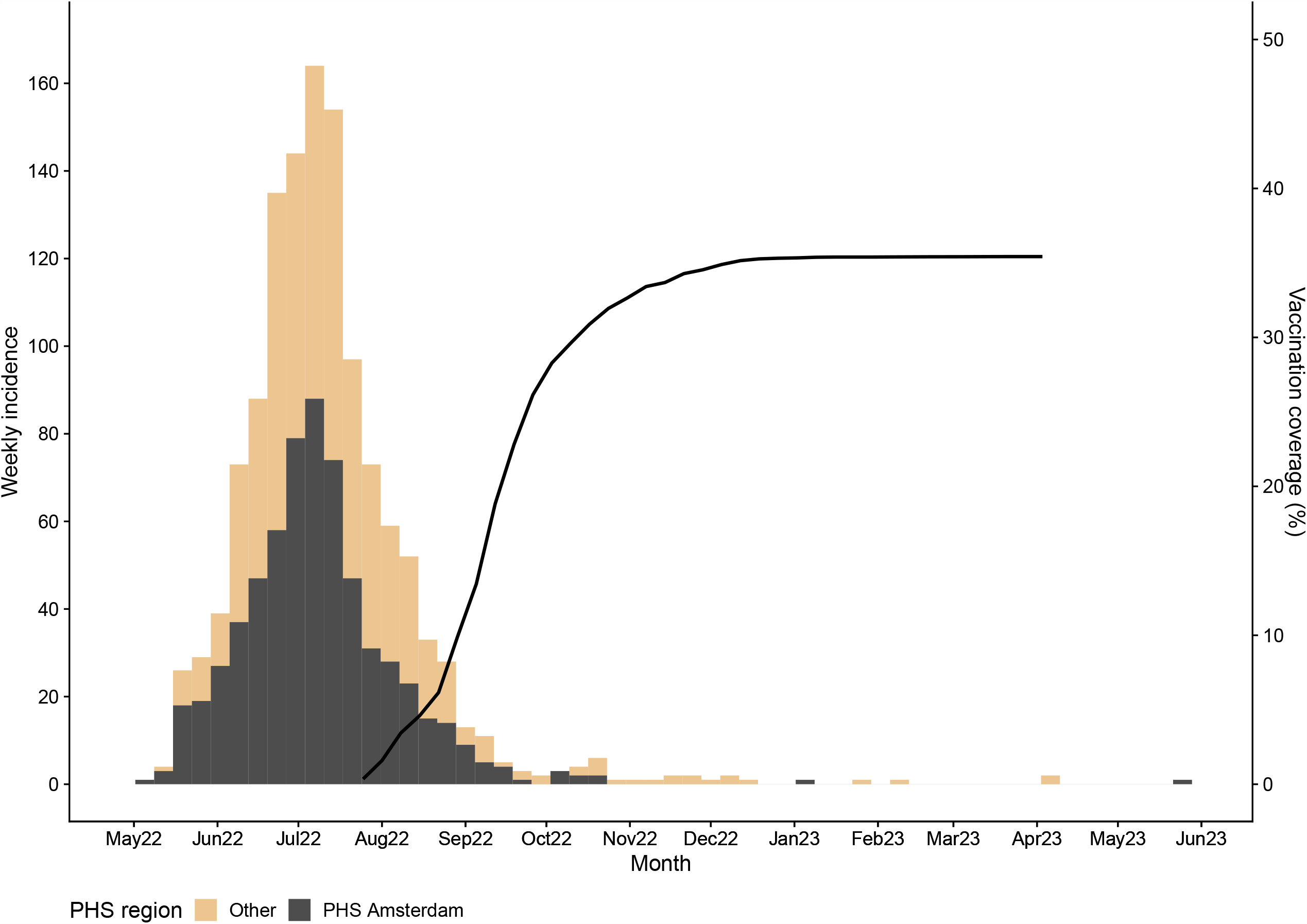
Epidemic curve by date of symptom onset and overall vaccination coverage. If the date of symptom onset was unknown, the earliest of either sampling date or notification date to the PHS was used. A bin width of 7 days was used and data were grouped by PHS Amsterdam (dark grey) versus the rest of the Netherlands (orange). Black line: vaccination coverage (fully vaccinated persons).

### Vaccine effectiveness

Between week 32 and 48 of 2022 (August 8 – December 4), a total of 181 individuals diagnosed with mpox were notified to the RIVM. Of those, 19 were excluded as they did not meet inclusion criteria to calculate the VE (i.e., not male, did not report to have sex with men, or vaccination status unknown). Table 3 shows the 162 included cases and VE per week. Overall VE of full vaccination against laboratory confirmed symptomatic mpox disease was estimated to be 68.2% (95% confidence interval 4.3% - 89.5%). The three fully vaccinated cases all received two PPV doses during the current vaccination programme.

**Table 3.** Cases with known vaccination status, vaccination coverage among the cases and population at risk, and VE (as estimated by the screening method) by week in August-November 2022.

| Week number | Start date of week | Cases |  |  | Population | Vaccine effectiveness |
| --- | --- | --- | --- | --- | --- | --- |
|  |  | Fully vaccinated cases | Total number of cases | Proportion of cases fully vaccinated | Proportion of population fully vaccinated |  |
| 32 | Aug 8 | 0 | 52 | 0.0% | 0.4% | 100% |
| 33 | Aug 15 | 0 | 31 | 0.0% | 1.6% | 100% |
| 34 | Aug 22 | 0 | 27 | 0.0% | 3.4% | 100% |
| 35 | Aug 29 | 0 | 13 | 0.0% | 4.6% | 100% |
| 36 | Sep 5 | 0 | 10 | 0.0% | 6.2% | 100% |
| 37 | Sep 12 | 0 | 6 | 0.0% | 9.9% | 100% |
| 38 | Sep 19 | 1 | 4 | 25.0% | 13.5% | -114.5% |
| 39 | Sep 26 | 0 | 0 | - | 18.8% | - |
| 40 | Oct 3 | 0 | 2 | 0.0% | 22.8% | 100% |
| 41 | Oct 10 | 1 | 5 | 20.0% | 26.1% | 29.4% |
| 42 | Oct 17 | 1 | 6 | 16.7% | 28.3% | 49.3% |
| 43 | Oct 24 | 0 | 2 | 0.0% | 29.6% | 100% |
| 44 | Oct 31 | 0 | 1 | 0.0% | 30.9% | 100% |
| 45 | Nov 7 | 0 | 0 | - | 31.9% | - |
| 46 | Nov 14 | 0 | 1 | 0.0% | 32.6% | 100% |
| 47 | Nov 21 | 0 | 1 | 0.0% | 33.4% | 100% |
| 48 | Nov 28 | 0 | 1 | 0.0% | 33.7% | 100% |
| **Overall** |  | **3** | **162** | **1.9%** |  | **68.2%** |

Cases: persons diagnosed with mpox
Population: all GBMSM and transgender persons invited for PPV

### Behavioural change

In 2022, 73,885 consultations were registered among GBMSM and transgender persons at SHC, of which 28,570 (38.7%) were HIV-PrEP-pilot consultations. The median age of included persons consulting SHC for HIV-PrEP was 37 years (IQR 30-49). In this group, Chi square analyses revealed a small but statistically significant decrease in the proportion of consultations where sex work in the past six months was reported in June versus May (2.4% vs. 3.3%, p = 0.049). Also, a significant increase in the proportion of consultations where group sex in the past six months was reported in August versus May (43% vs. 38%, p < 0.001) (see Supplementary Files). No trend was seen in the other indicators.

## Discussion

After the first detection of mpox outside of endemic regions in May 2022, a large global outbreak occurred among GBMSM with a peak in incidence in the summer of 2022. After the peak, the incidence dropped rapidly, both in the Netherlands and in most of the other non-endemic affected areas.^8,18^ In addition to a targeted risk communication campaign, the Netherlands rapidly introduced control measures including PEP for contacts and a targeted PPV programme to protect those at highest risk of mpox. This resulted in an overall vaccination coverage of 35.4% in the target group by April 30, 2023, with differences between the different indication groups. In addition, a VE of full vaccination against laboratory confirmed symptomatic mpox disease of 68.2% was found, which is comparable to estimates from other studies. For one MVA-BN dose, VE estimates range from 36%-86%,^19–23^ and for two doses from 66% to 89%.^21–23^ Persons who received a childhood smallpox vaccination were also included in the referenced studies. Those persons would have been considered fully vaccinated after one MVA-BN vaccination in our definition. Therefore, it was to be expected that our estimated VE is a bit lower than the estimate for two doses MVA-BN.

The impact of the vaccination programme on the course of the epidemic is difficult to establish. Generally, a good immune response is seen two weeks after the first dose of MVA-BN, with seroconversion seen in 91% of subjects in a study by Pittman et al.^24^ However, antibody titres increased significantly after a second vaccination with MVA-BN, given four weeks after the first vaccination. When also taking an incubation period of 8 to 9 days into account,^25,26^ it may take several days to weeks before a significant effect of the vaccination programme on infections could be noticed. In addition, the timing and speed with which the vaccination programme is implemented also influences its impact. In the Netherlands, the PPV programme started after the peak of the outbreak and by the time PPV would be effective, the incidence was already down to about 5 cases per day, making it unlikely that PPV contributed significantly to the decline of the outbreak.

Unfortunately, the effect of PEP given to high-risk contacts of mpox cases cannot be assessed, as national data are lacking. PEP is unlikely to have contributed substantially given that the incubation period of mpox is about equal to, or possibly even shorter than the time to generate vaccine-induced immunity.^24,25^ Furthermore, as it takes time to identify high-risk contacts of a mpox case, invite them for vaccination and administer the vaccine, the impact of PEP to prevent mpox after exposure is likely to be negligible. However, PEP vaccinations may have contributed to the overall effect of mpox vaccination, as they were administered to the persons at highest risk for mpox infection.

Correct calculation of the uptake, vaccination coverage and assessing VE was challenging due to limitations regarding the quality of data and lack of data on denominators. Identifying and personally inviting the population at risk through multiple health care providers was challenging. In addition, some people used an alias, and privacy guidelines prohibited direct comparison of persons possibly invited multiple times. In an online survey conducted from July 1^st^ to mid-August 2023, 16.3% of the GBMSM invited for vaccination reported to have received more than one invitation for vaccination (Adam P, unpublished). Also, exact numbers of invitations sent out by GPs and HIV health care providers were not known. Finally, it is known that several HIV health care providers invited all their patients instead of only those at highest risk of mpox, which could explain the lower PPV uptake in this group. Overall, the denominator is likely overestimated, but it is unknown to what extent. On the other hand, the number of administered vaccinations might be underestimated, since not all data were available due to privacy constraints. Altogether, we believe we may have underestimated the uptake, vaccination coverage and VE. Thus, our results emphasise the importance of a vaccination register with national coverage and adequate privacy assurance, to facilitate adequate monitoring and evaluation of future vaccination programmes in outbreak settings.

Our observations regarding sexual activity differ from reports from external sources. A behavioural survey conducted in August 2022 among 2,460 GBMSM in the Netherlands (Adam P, unpublished), indicated that half (50.4%) of the respondents had reduced their number of partners since the start of the outbreak, and two-thirds (65.5%) had avoided sex on premises venues. Similar behavioural adaptations to mpox were observed among GBMSM in the US as well.^27^ However, when examining our data on SHC visits, we could not identify comparable changes in behaviour, nor in positivity rates of STI. Most behavioural indicator variables remained relatively stable in 2022. The changes observed simply entailed a minor reduction in reports of sex work in the past six months in June compared to May, and a slight increase in reports of group sex in the past six months in August versus May. Although these findings align with the hypothesis of decreased sexual activity during the outbreak followed by a reengagement in sexual activity as the outbreak waned, the extent of these changes observed in our data was extremely limited.

The fact that, at each point in time, our behavioural indicator variables consisted of reports for the past six months (rather than the last month, for instance) could have hindered the detection of more significant changes in behaviour, particularly if GBMSM’s behavioural adaptations primarily occurred during the brief period when high numbers of mpox cases were reported. Also, HIV-PrEP users who regularly visit SHC on which the analyses were based may not be representative of the entire population of GBMSM at risk for mpox.

Based on the above findings, a likely explanation for the declining course of the outbreak may be the impact of acquired immunity, either by natural infection or PEP, in a substantial proportion of those most at risk. This is also in line with results from several modelling studies that conclude that immunity due to infection in the most sexually active GBMSM may be the main driver of the decline.^28–31^ Modelling results using data on Dutch mpox cases before the start of the PPV programme indicate that increased immunity due to infections may have limited further growth of the mpox epidemic, but could not exclusively explain the fast decline after the peak. This suggests that behavioural adaptations among GBMSM may have accelerated the decrease observed before the start of PPV.^30^ It may also explain the observed vaccine uptake, which was notably lower than expected based on previous studies investigating vaccine acceptance among GBMSM in the Netherlands. They found that 70-85% of GBMSM in July-September 2022 were willing to accept mpox vaccination.^32,33^ The perceived sense of urgency may have been low at the time people were invited for PPV, especially for the second dose, as the outbreak was already declining. This may have led to a lower vaccine uptake and vaccination coverage than expected.

Although we hypothesise that it is unlikely that PPV was the main driver of decline of the mpox outbreak, vaccination might play an important role to prevent future outbreaks, especially since the VE is expected to be relatively high. Fast implementation of future vaccination programmes is essential to be effective in preventing new cases.^18^ Immunity due to infection and vaccination among the most sexually active GBMSM reduces the risk of resurgence of mpox^28–30^ and will protect against (severe) disease. The vaccination programme in the Netherlands continued in 2023 and took newly identified risk factors, such as visiting sex venues, into account.^34,35^ Continued vaccination programmes are needed to offer vaccination to new people meeting the criteria for vaccination or recently arrived migrant GBMSM. Surveillance of mpox and evaluation of the vaccination programmes, including assessment of VE, are of critical importance to inform future mpox prevention and control.

## Data Availability

Deidentified individual participant data included in this study will not be made available to others because of the sensitive nature of participant data extracted from participating public health services.

## Contributions

SJMH and MRH conceptualized the study. SJMH, MRH, ELMOdC, IJMW, CEvE designed the study. MRH, IJMW, CEvE, SJL, PJJ, CvB, GGCvR, EH collected the data. MRH, IJMW, CEvE did the statistical analyses. MRH and IJMW wrote the first draft of the manuscript. All authors contributed to writing the manuscript, and all approved the final version.

## Acknowledgments

We thank Maartje Visser from the RIVM for selecting the persons belonging to the indication groups from the national database. We also thank Birgit van Benthem, Susan van den Hof and Maria Xiridou from the RIVM for their critical view and suggestions for improvement of the manuscript. Furthermore, we thank all the PHS and SHC in the Netherlands for their commitment during the mpox outbreak and PPV programme.

This work was supported by the Dutch Ministry of Health, Welfare and Sport.

## Disclaimer

CvE is a fellow of the ECDC Fellowship Programme, supported financially by the European Centre for Disease Prevention and Control The views and opinions expressed herein do not state or reflect those of ECDC. ECDC is not responsible for the data and information collation and analysis and cannot be held liable for conclusions or opinions drawn.

## References

1. Angelo KM, Smith T, Camprubi-Ferrer D, Balerdi-Sarasola L, Diaz Menendez M, Servera-Negre G, et al. Epidemiological and clinical characteristics of patients with monkeypox in the GeoSentinel Network: a cross-sectional study. Lancet Infect Dis 2022; 2: 196–206. DOI: 10.1016/S1473-3099(22)00651-X.

2. McCollum AM, Shelus V, Hill A, Traore T, Onoja B, Nakazawa Y, et al. Epidemiology of Human Mpox - Worldwide, 2018-2021. MMWR Morb Mortal Wkly Rep 2023; 72: 68–72. DOI: 10.15585/mmwr.mm7203a4.

3. Di Giulio DB, Eckburg PB. Human monkeypox: an emerging zoonosis. Lancet Infect Dis 2004;4: 15–25. DOI: 10.1016/s1473-3099(03)00856-9.

4. Centers for Disease Control and Prevention (CDC). Multistate outbreak of monkeypox--Illinois, Indiana, and Wisconsin, 2003. MMWR Morb Mortal Wkly Rep 2003; 52: 537–40.

5. Gross E. Update on emerging infections: news from the Centers for Disease Control and prevention. Update: Multistate outbreak of monkeypox--Illinois, Indiana, Kansas, Missouri, Ohio, and Wisconsin, 2003. Ann Emerg Med 2003; 42: 660–2. DOI: 10.1016/S0196064403008199.

6. Vaughan AM, Cenciarelli O, Colombe S, Alves de Sousa L, Fischer N, Gossner C, et al. A large multi-country outbreak of monkeypox across 41 countries in the WHO European Region, 7 March to 23 August 2022. Euro Surveill 2022; 27: pii=2200620. DOI: 10.2807/1560-7917.ES.2022.27.36.2200620.

7. Laurenson-Schafer H, Sklenovska N, Hoxha A, Kerr SM, Ndumbi P, Fitzner J, et al. Description of the first global outbreak of mpox: an analysis of global surveillance data. Lancet Glob Health 2023; 11: e1012–23. DOI: 10.1016/S2214-109X(23)00198-5.

8. World Health Organization (WHO). 2022-23 Mpox (Monkeypox) Outbreak: Global Trends. Available via https://worldhealthorg.shinyapps.io/mpx_global/ (accessed August 1, 2023).

9. van Ewijk CE, Miura F, van Rijckevorsel G, de Vries HJC, Welkers MRA, van den Berg OE, et al. Mpox outbreak in the Netherlands, 2022: public health response, characteristics of the first 1,000 cases and protection of the first-generation smallpox vaccine. Euro Surveill 2023; 28: pii=2200772. DOI: 10.2807/1560-7917.ES.2023.28.12.2200772.

10. European Medicines Agency (EMA). Imvanex. Available via https://www.ema.europa.eu/en/medicines/human/EPAR/imvanex (accessed August 1, 2023).

11. Dutch National Institute for Public Health and the Environment (RIVM). Mpoxvaccinatie Uitvoeringsrichtlijn [Mpox vaccination practice guideline]. Available via https://lci.rivm.nl/richtlijnen/monkeypoxvaccinatie (accessed October 1, 2022).

12. Ministry of Health, Welfare and Sport. Factsheet Pokken [Factsheet Smallpox], version March 3, 2003.

13. Taube JC, Rest EC, Lloyd-Smith JO, Bansal S. The global landscape of smallpox vaccination history and implications for current and future orthopoxvirus susceptibility: a modelling study. Lancet Infect Dis 2023; 23: 454–62. DOI: 10.1016/S1473-3099(22)00664-8.

14. Soa Aids Nederland. Uitbreiding voorlichtingscampagne mpox [Expansion of mpox awareness campaign]. Available via https://www.soaaids.nl/nl/professionals/actueel/nieuwsbericht/uitbreiding-voorlichtingscampagne-mpox (accessed August 1, 2023).

15. Minister of Health, Welfare and Sport. Regeling apenpokken [Monkeypox regulation]. Available via https://wetten.overheid.nl/BWBR0046692/2022-05-21/0/informatie (accessed October 1, 2022).

16. Farrington CP. Estimation of vaccine effectiveness using the screening method. Int J Epidemiol 2023; 52: 14–21. DOI: 10.1093/ije/dyac207.

17. Flannery B, Andrews N, Feikin D, Patel MK. Commentary: Estimation of vaccine effectiveness using the screening method. Int J Epidemiol 2023; 52: 19–21. DOI: 10.1093/ije/dyac013.

18. Vairo F, Leone S, Mazzotta V, Piselli P, De Carli G, Lanini S. The possible effect of sociobehavioral factors and public health actions on the mpox epidemic slowdown. Int J Infect Dis 2023; 130: 83–5. DOI: 10.1016/j.ijid.2023.03.005.

19. Bertran M, Andrews N, Davison C, Dugbazah B, Boateng J, Lunt R, et al. Effectiveness of one dose of MVA-BN smallpox vaccine against mpox in England using the case-coverage method: an observational study. Lancet Infect Dis 2023; 23: 828–35. DOI: 10.1016/S1473-3099(23)00057-9.

20. Wolff Sagy Y, Zucker R, Hammerman A, Markovits H, Gur Arieh N, Abu Ahmad W, et al. Realworld effectiveness of a single dose of mpox vaccine in males. Nat Med 2023; 29: 748–52. DOI: 10.1038/s41591-023-02229-3.

21. Deputy NP, Deckert J, Chard AN, Sandberg N, Moulia DL, Barkley E, et al. Vaccine Effectiveness of JYNNEOS against Mpox Disease in the United States. N Engl J Med 2023; 388: 2434–43. DOI: 10.1056/NEJMoa2215201.

22. Dalton AF, Diallo AO, Chard AN, Moulia DL, Deputy NP, Fothergill A, et al. Estimated Effectiveness of JYNNEOS Vaccine in Preventing Mpox: A Multijurisdictional Case-Control Study - United States, August 19, 2022-March 31, 2023. MMWR Morb Mortal Wkly Rep 2023; 72: 553–8. 20230519. DOI: 10.15585/mmwr.mm7220a3.

23. Rosenberg ES, Dorabawila V, Hart-Malloy R, Anderson BJ, Miranda W, O’Donnell T, et al. Effectiveness of JYNNEOS Vaccine Against Diagnosed Mpox Infection - New York, 2022. MMWR Morb Mortal Wkly Rep 2023; 72: 559–63. DOI: 10.15585/mmwr.mm7220a4.

24. Pittman PR, Hahn M, Lee HS, Koca C, Samy N, Schmidt D, et al. Phase 3 Efficacy Trial of Modified Vaccinia Ankara as a Vaccine against Smallpox. N Engl J Med 2019; 381: 1897–908. DOI: 10.1056/NEJMoa1817307.

25. Miura F, van Ewijk CE, Backer JA, Xiridou M, Franz E, Op de Coul E, et al. Estimated incubation period for monkeypox cases confirmed in the Netherlands, May 2022. Euro Surveill 2022; 27: pii=2200448. DOI: 10.2807/1560-7917.ES.2022.27.24.2200448.

26. McFarland SE, Marcus U, Hemmers L, Miura F, Inigo Martinez J, Martin Martinez F, et al. Estimated incubation period distributions of mpox using cases from two international European festivals and outbreaks in a club in Berlin, May to June 2022. Euro Surveill 2023; 28: pii=2200806. DOI: 10.2807/1560-7917.ES.2023.28.27.2200806.

27. Delaney KP, Sanchez T, Hannah M, Winslow Edwards O, Carpino T, Agnew-Brune C, et al. Strategies Adopted by Gay, Bisexual, and Other Men Who Have Sex with Men to Prevent Monkeypox virus Transmission - United States, August 2022. MMWR Morb Mortal Wkly Rep 2022; 71: 1126–30. DOI: 10.15585/mmwr.mm7135e1.

28. Kupferschmidt K. Monkeypox outbreak is ebbing-but why exactly?Science 2022; 378: 343. DOI: 10.1126/science.adf4961.

29. Brand SPC, Cavallaro M, Cumming F, Turner C, Florence I, Blomquist P, et al. The role of vaccination and public awareness in forecasts of Mpox incidence in the United Kingdom. Nat Commun 2023; 14: 4100. DOI: 10.1038/s41467-023-38816-8.

30. [preprint] Xiridou M, Miura F, Adam PCG, Op de Coul E, de Wit J, Wallinga J. The fading of the mpox outbreak among men who have sex with men: a mathematical modelling study. medRxiv 2023. DOI: 10.1101/2023.01.31.23285294.

31. Murayama H, Pearson CAB, Abbott S, Miura F, Jung S, Fearon E, et al. Accumulation of immunity in heavy-tailed sexual contact networks shapes mpox outbreak sizes. J Infect Dis 2023; jiad254. DOI: 10.1093/infdis/jiad254.

32. Wang H, d’Abreu de Paulo KJI, Gultzow T, Zimmermann HML, Jonas KJ. Monkeypox self-diagnosis abilities, determinants of vaccination and self-isolation intention after diagnosis among MSM, the Netherlands, July 2022. Euro Surveill 2022; 27: pii=2200603. DOI: 10.2807/1560-7917.ES.2022.27.33.2200603.

33. Dukers-Muijrers N, Evers Y, Widdershoven V, Davidovich U, Adam PCG, Op de Coul E, et al. Mpox vaccination willingness, determinants, and communication needs in gay, bisexual, and other men who have sex with men, in the context of limited vaccine availability in the Netherlands (Dutch Mpox-survey). Front Public Health 2022; 10: 1058807. DOI: 10.3389/fpubh.2022.1058807.

34. [preprint] Adam PCG, Op de Coul ELM, Zantkuijl P, Bos H, Xiridou M, Blom C, et al. An assessment of rates and covariates of mpox diagnosis and vaccination provides evidence to refine eligibility criteria for mpox vaccination among gay, bisexual and other men who have sex with men in the Netherlands. medRxiv 2023. DOI: 10.1101/2023.02.28.23286578.

35. Dutch National Institute for Public Health and the Environment (RIVM). Mpoxvaccinatie Uitvoeringsrichtlijn [Mpox vaccination practice guideline]. Available via https://lci.rivm.nl/richtlijnen/monkeypoxvaccinatie (accessed August 1, 2023).

