## Supplementary Material for "Factors that influenced the course of the mpox outbreak in the Netherlands, 2022-2023"

**Supplementary Information**

**Mpox mandatory notification**

Mpox was declared a notifiable disease in group A on May 20, 2022.[1] A group A status includes direct mandatory notification of suspected and confirmed cases by treating physicians and laboratories to the Public Health Services (PHS), enabling timely infection control measures, such isolation, quarantine and PEP vaccination. On December 3, 2022 the notification status was scaled down to group B1, which entails case isolation and quarantine advice (not enforceable) coupled with mandatory notification of only laboratory confirmed cases; the reporting time was changed to within one workday.

**Mpox PEP and PPV vaccination programme the Netherlands**

PEP vaccination with MVA-BN started immediately after the confirmation of the first case in the Netherlands on May 20, 2022. PEP was given preferably within 4, but up to 14 days after the first exposure. The definition for high- and medium-risk contacts was described in a national guideline.[2] One dose was offered, except for persons with continued substantial risk for exposure to MPXV, for example if they had multiple varying sexual partners. They were offered a second dose after four weeks. Persons who received a (first-generation) childhood smallpox vaccination were considered protected after one MVA-BN vaccination. An exception were immunocompromised patients (CD4 number <200 and/or another immunocompromising condition), who should receive two MVA-BN vaccinations, regardless of childhood smallpox vaccination.[2]

To control the outbreak of mpox in the Netherlands, also a PPV vaccination programme was set up.[2] Due to limited availability of the vaccine, only persons at high risk were offered vaccination. Indication groups were defined by expert opinion on June 22, 2022. These were: 1a. GBMSM and transgender persons using HIV-pre-exposure prophylaxis (PrEP) via Sexual Health Centres (SHC); 1b. GBMSM and transgender persons on the waiting list for the HIV-PrEP-pilot via SHC; 1c. GBMSM and transgender persons using HIV-PrEP via their general practitioner (GP); 2. GBMSM and transgender persons living with HIV who are being screened for hepatitis C as a proxy for high-risk behaviour; 3. Other GBMSM and transgender persons at high-risk, defined as visiting a SHC in the previous six months because they were notified for a sexually transmitted infection (STI) and/or HIV by a sex partner; because of a syphilis, gonorrhoea or chlamydia infection; or those who reported more than three sex partners in the past six months.[2,3]

In the Netherlands, all visits to SHC are available in a national surveillance database (SOAP) at the RIVM. This database contains pseudonymised data with, amongst others, information on age, sex, (sexual) behaviour, STI diagnoses, HIV status, and use of HIV-PrEP (a national PrEP pilot programme has been implemented at SHC, in which HIV-PrEP is provided to individuals who are at a higher risk to acquire HIV). Based on the SOAP database, combined with estimates on the number of GBMSM and transgender persons living with HIV, the number of GBMSM and transgender persons using HIV-PrEP via a GP, and the number of GBMSM and transgender persons on the waiting list for the HIV-PrEP-pilot, it was initially estimated that about 33,000 persons were eligible for vaccination.

Selection of persons belonging to the indication groups was done in several ways. The RIVM selected individuals from the SOAP database that met the high-risk criteria (mostly indication groups 1a and 3), deduplicated the list and distributed a regional list to each SHC. There, the pseudonymised patient numbers could be used to invite people for vaccination. About half of the SHC used this list, the other half made their own selection from their local database. The national database does not contain information of those on the waiting list for the HIV-PrEP-pilot (indication group 1b), thus the

SHC had to extract this from their own database. For indication group 1c (i.e., persons using HIV-PrEP via their GP) PHS were asked to contact all GPs in their region that prescribe HIV-PrEP and ask them to invite the respective patients for PPV. A similar approach was adopted for persons living with HIV (indication group 2), where HIV health care providers were asked by the PHS to invite their patients that met the criteria for vaccination.

Administered PPV vaccinations were registered by the PHS in either of two systems: a reporting system hosted by the RIVM called 'Osiris', used by 4 PHS; or a vaccination register developed especially for mpox vaccination called 'iMPeX', used by 21 PHS. These data were combined at the RIVM, resulting in a database with national coverage.

**References**


1. Minister of Health, Welfare and Sports. Regeling apenpokken [Monkeypox regulation]. Available via https://wetten.overheid.nl/BWBR0046692/2022-05-21/0/informatie (accessed October 1, 2022).
2. Dutch National Institute for Public Health and the Evironment. Mpoxvaccinatie Uitvoeringsrichtlijn [Mpox vaccination practice guideline]. Available via https://lci.rivm.nl/richtlijnen/monkeypoxvaccinatie (accessed October 1, 2022).
3. Adviesbrief Deskundigenberaad monkeypox [Advice of the expert panel on monkeypox], June 30, 2022. Available via https://open.overheid.nl/Details/ronl-d19aa3f1530c221f0a9a0041ab82773a95b4e9f6/1.

### Supplementary Figures

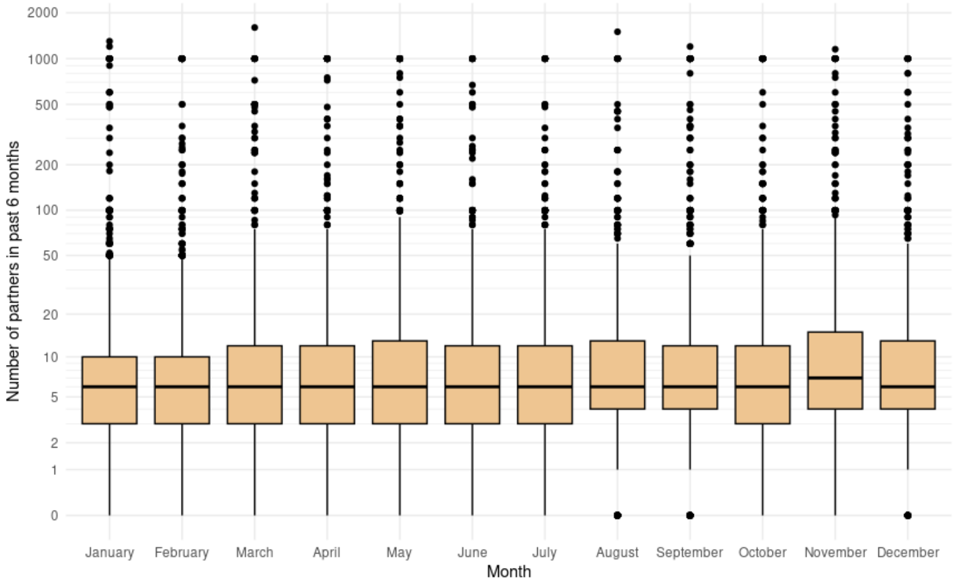


**Figure S1. Boxplot of number of sex partners in the past 6 months reported in GBMSM and transgender persons using HIV-PrEP, 2022 (vertical axis log transformed)**

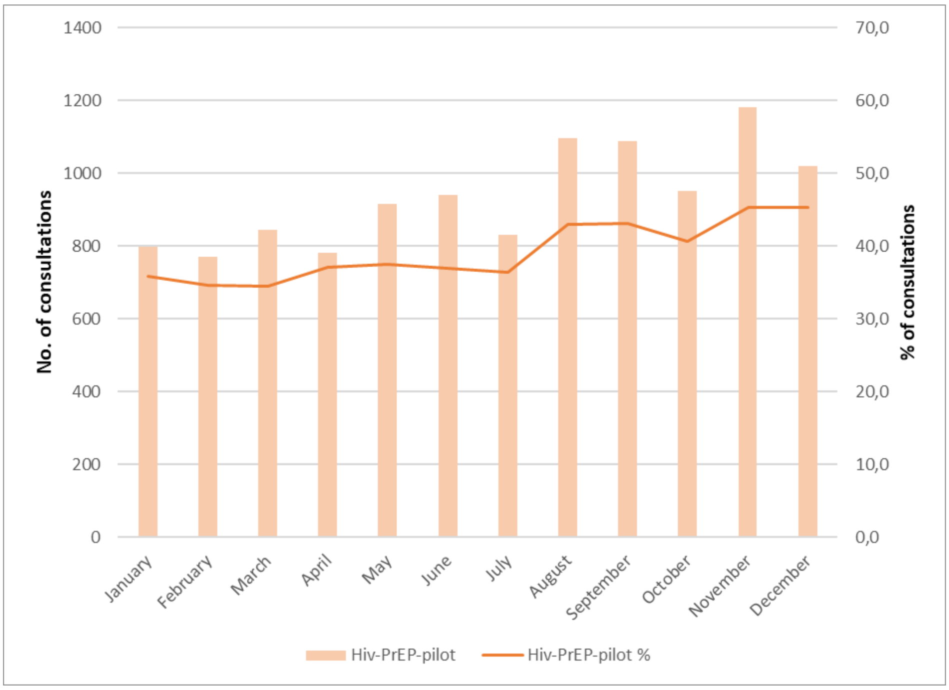


**Figure S2. Number and percentage of consultations where group sex in the past 6 months is reported in GBMSM and transgender persons using HIV-PrEP, 2022**

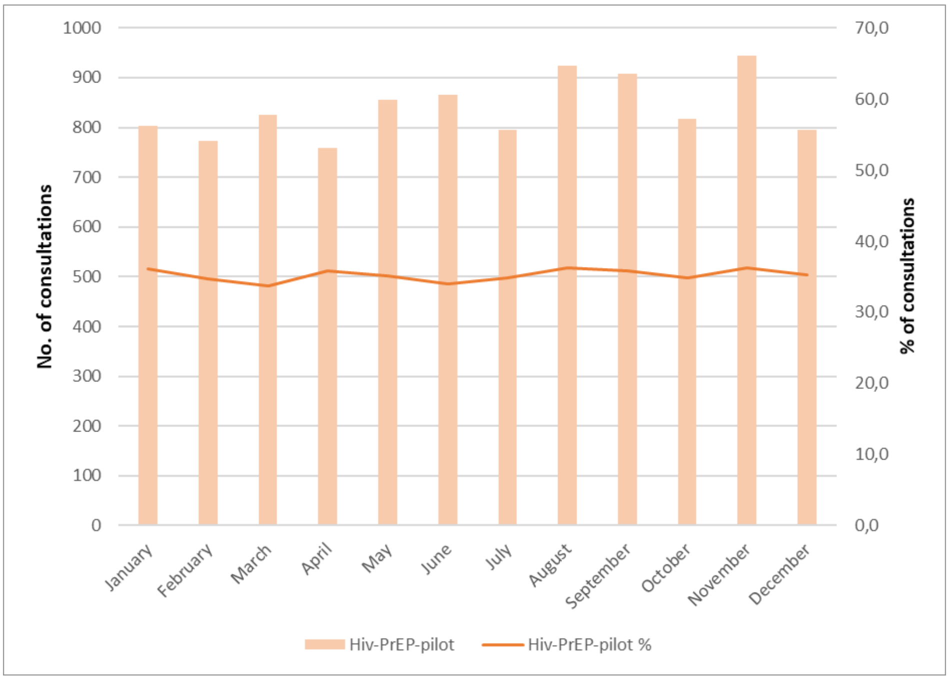


**Figure S3. Number and percentage of consultations where using drugs before or during sex (chemsex) in the past 6 months is reported in GBMSM and transgender persons using HIV-PrEP, 2022**

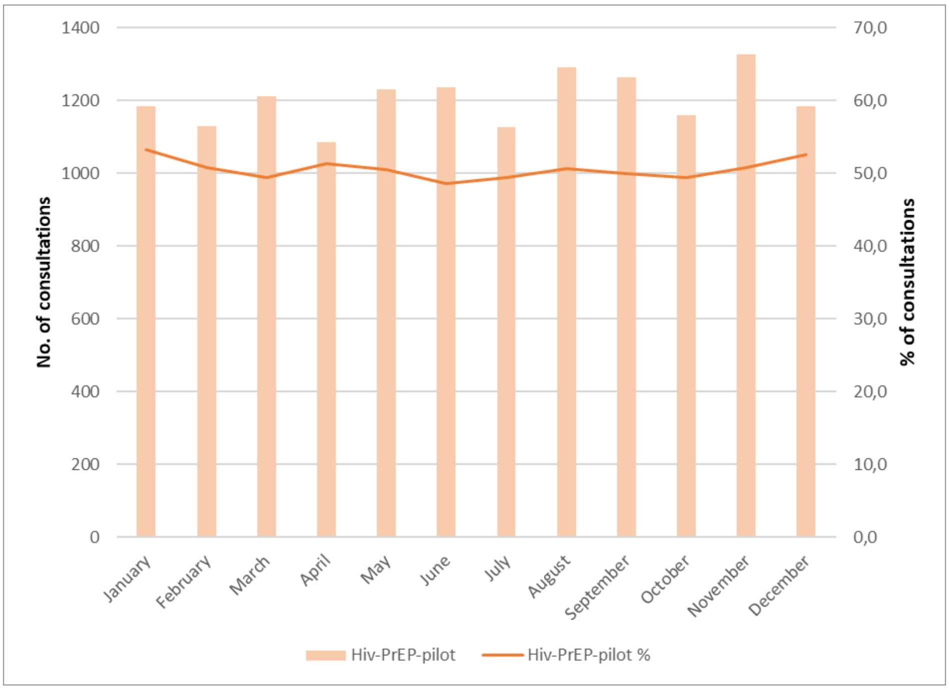


**Figure S4. Number and percentage of consultations where condomless anal sex (always condomless) in the past 6 months is reported in GBMSM and transgender persons using HIV-PrEP, 2022**

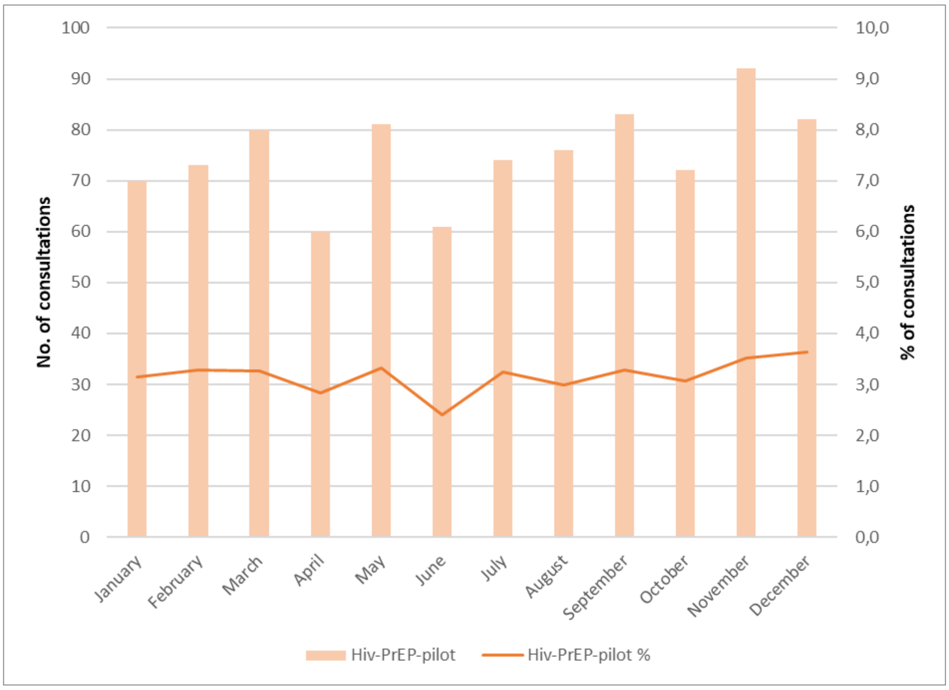


**Figure S5. Number and percentage of consultations where sex work in the past 6 months is reported in GBMSM and transgender persons using HIV-PrEP, 2022**

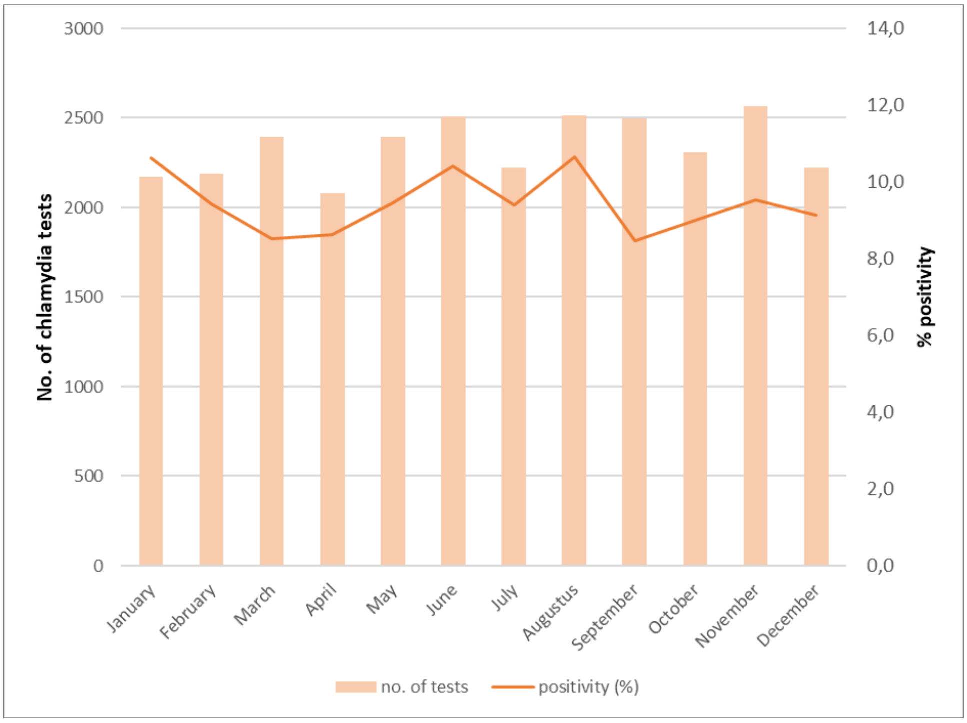


**Figure S6. Number of chlamydia tests and chlamydia positivity in GBMSM and transgender persons using HIV-PrEP, 2022**

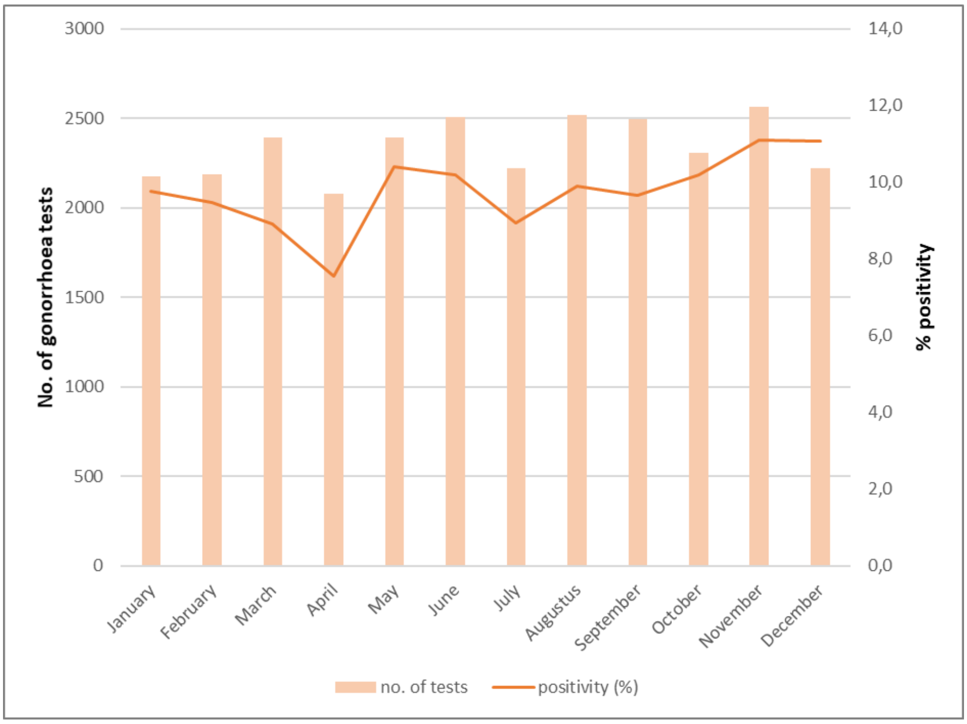


**Figure S7. Number of gonorrhoea tests and gonorrhoea positivity in GBMSM and transgender persons using HIV-PrEP, 2022**

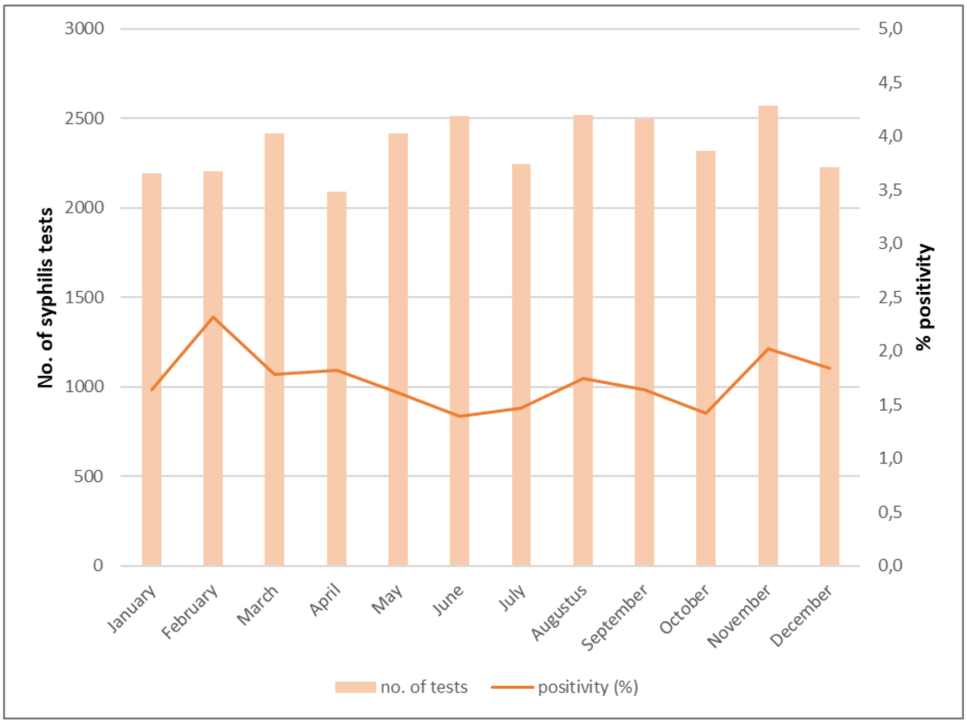


**Figure S8. Number of syphilis tests and infectious syphilis positivity in GBMSM and transgender persons using HIV-PrEP, 2022**

**Supplementary Tables**

**Table S1. Mann-Whitney U test comparing the median number of sex partners in the past 6 months reported during visits in the outbreak months (June, July and August 2022) to pre-outbreak (May 2022) in GBMSM and transgender persons using HIV-PrEP**

| **Month** | **Median** | **Interquartile range** | **p-value** |
|---|---|---|---|
| May | 6 | 3 – 13 | Ref |
| June | 6 | 3 – 12 | 0.672 |
| July | 6 | 3 – 12 | 0.889 |
| August | 6 | 4 – 13 | 0.264 |

**Table S2. Chi square analyses comparing the percentage of consultations where group sex in the past 6 months is reported during visits in the outbreak months (June, July and August 2022) to pre-outbreak (May 2022) in GBMSM and transgender persons using HIV-PrEP**

| **Month** | **% of consultations** | **p-value** |
|---|---|---|
| May | 37.6% | Ref |
| June | 36.9% | 0.640 |
| July | 36.4% | 0.414 |
| August | 43.0% | **<0.001** |

**Table S3. Chi square analyses comparing the percentage of consultations where using drugs before or during sex (chemsex) in the past 6 months is reported during visits in the outbreak months (June, July and August 2022) to pre-outbreak (May 2022) in GBMSM and transgender persons using HIV-PrEP**

| **Month** | **% of consultations** | **p-value** |
|---|---|---|
| May | 35.2% | Ref |
| June | 34.0% | 0.387 |
| July | 34.9% | 0.827 |
| August | 36.3% | 0.398 |

**Table S4. Chi square analyses comparing the percentage of consultations where condomless anal sex in the past 6 months is reported during visits in the outbreak months (June, July and August 2022) to pre-outbreak (May 2022) in GBMSM and transgender persons using HIV-PrEP**

| **Month** | **% of consultations** | **p-value** |
|---|---|---|
| May | 50.6% | Ref |
| June | 48.6% | 0.169 |
| July | 49.4% | 0.407 |
| August | 50.7% | 0.938 |

**Table S5. Chi square analyses comparing the percentage of consultations where sex work in the past 6 months is reported during visits in the outbreak months (June, July and August 2022) to pre-outbreak (May 2022) in GBMSM and transgender persons using HIV-PrEP**

| Month | % of consultations | p-value |
|---|---|---|
| May | 3.3% | Ref |
| June | 2.4% | **0.049** |
| July | 3.2% | 0.868 |
| August | 3.0% | 0.487 |

**Table S6. Chi square analyses comparing chlamydia positivity in the outbreak months (June, July and August 2022) to pre-outbreak (May 2022) in GBMSM and transgender persons using HIV-PrEP**

| Month | % positivity | p-value |
|---|---|---|
| May | 9.4% | Ref |
| June | 10.4% | 0.262 |
| July | 4.4% | 0.965 |
| August | 10.6% | 0.161 |

**Table S7. Chi square analyses comparing gonorrhoea positivity in the outbreak months (June, July and August 2022) to pre-outbreak (May 2022) in GBMSM and transgender persons using HIV-PrEP**

| Month | % positivity | p-value |
|---|---|---|
| May | 10.4% | Ref |
| June | 10.2% | 0.824 |
| July | 8.9% | 0.096 |
| August | 9.9% | 0.559 |

**Table S8. Chi square analyses comparing infectious syphilis positivity in the outbreak months (June, July and August 2022) to pre-outbreak (May 2022) in GBMSM and transgender persons using HIV-PrEP**

| Month | % positivity | p-value |
|---|---|---|
| May | 1.6% | Ref |
| June | 1.4% | 0.262 |
| July | 1.5% | 0.965 |
| August | 1.7% | 0.161 |
